# Sex and aging signatures of proteomics in human cerebrospinal fluid identify distinct clusters linked to neurodegeneration

**DOI:** 10.1101/2024.06.18.24309102

**Authors:** Dahun Seo, Cheolmin Matthew Lee, Catherine Apio, Gyujin Heo, Jigyasha Timsina, Pat Kohlfeld, Merce Boada, Adelina Orellana, Maria Victoria Fernandez, Agustin Ruiz, John C. Morris, Suzanne E. Schindler, Taesung Park, Carlos Cruchaga, Yun Ju Sung

**Author notes:** Corresponding Author: Yun Ju Sung, PhD Washington University, School of Medicine 4444 Forest Park Ave, Room 5501, Box 8134 St. Louis, MO 63108. These authors contributed equally (Dahun Seo, Cheolmin Matthew Lee, Catherine Apio). These authors contributed equally (Carlos Cruchaga, Yun Ju Sung).

## Abstract

Sex and age are major risk factors for chronic diseases. Recent studies examining age-related molecular changes in plasma provided insights into age-related disease biology. Cerebrospinal fluid (CSF) proteomics can provide additional insights into brain aging and neurodegeneration. By comprehensively examining 7,006 aptamers targeting 6,139 proteins in CSF obtained from 660 healthy individuals aged from 43 to 91 years old, we subsequently identified significant sex and aging effects on 5,097 aptamers in CSF. Many of these effects on CSF proteins had different magnitude or even opposite direction as those on plasma proteins, indicating distinctive CSF-specific signatures. Network analysis of these CSF proteins revealed not only modules associated with healthy aging but also modules showing sex differences. Through subsequent analyses, several modules were highlighted for their proteins implicated in specific diseases. Module 2 and 6 were enriched for many aging diseases including those in the circulatory systems, immune mechanisms, and neurodegeneration. Together, our findings fill a gap of current aging research and provide mechanistic understanding of proteomic changes in CSF during a healthy lifespan and insights for brain aging and diseases.

## Introduction

Aging is a complex and gradual process involving both physiological and pathological changes characterized by progressive functional decline, resulting in decreased fitness, increased susceptibility, and vulnerability to diseases, and eventually death^1^. Many chronic diseases, including obesity, diabetes, cardiovascular disease (CVD)^2^, stroke, and neurodegenerative brain diseases,^3^ occur more frequently as aging, and multiple of these diseases often co-occur. These age-related diseases are the leading causes of mortality and increasing healthcare costs. With increased life expectancy, these costs are continually rising and represent an ever-growing challenge. In addition to aging, clear sex differences exist in many of these diseases and continue to present across the lifespan. Parkinson’s disease (PD)^4^ and cardiovascular diseases^5^ are more prevalent in males, whereas Alzheimer’s disease (AD) is more prevalent in females^6^. Other diseases including osteoporosis^7^ and multiple sclerosis^8^ also show the disparity between males and females. Therefore, understanding the mechanisms of aging and sex differences and their association with age-related diseases and resulting comorbidity conditions is essential for developing therapies that mitigate age-related disease and prolong healthy aging and longevity.^9–12^

Several studies using omics data including methylations, transcriptomics, and proteomics^13^ examined age-related changes, identifying a broad spectrum of biomarkers associated with the aging process and providing new insights into the biology of chronic diseases^14^. Compared to methylation and transcriptomic levels, proteins are final effectors of most physiological pathways, often dysregulated in diseases, and important drug targets. Most human proteomic studies have considered plasma due to easy accessibility^13,15^. For example, Lehallier et al. identified dynamic changes in several plasma proteomes throughout the life span, demonstrating biological aging is a better predictor for morbidity and mortality than chronological aging^16,17^. Koichiro et al. identified sex differences in serum protein composition^18^. Couillard et al. found that females have three times higher plasma leptin and weight-losing hormones than males^19^. These and several other plasma proteomic studies have successfully utilized plasma proteome profiling, elucidated the aging process, and provided quantitative and functional characterization of pathological conditions^20^. Compared to plasma, cerebrospinal fluid (CSF) is adjacent to the brain and can better capture the aging process of brain and central nervous system. For most neurodegenerative diseases, CSF proteins are gold standard biomarkers and provide critical pathological changes occurring in the brain. As certain age-associated diseases including neurodegeneration demonstrate their pathological evidence through CSF, studying CSF aging signatures has immense potential in uncovering idiopathic conditions that cannot be discovered using other specimens. However, due to the perceived invasiveness of obtaining CSF samples via lumbar puncture, relatively few studies collect CSF samples from healthy individuals.

We hypothesize that the CSF proteome has different sex and aging signatures when compared to the plasma proteome, which in turn can uncover biological aging processes that may be specific to brain aging. To test this hypothesis, we generated 7,006 aptamers targeting 6,162 proteins in CSF from 994 cognitively normal individuals across three large cohorts (Table 1). We pursued an unbiased and hypothesis-free approach to examine the impact of aging and sex differences on CSF proteome in the discovery cohort and validated them in the two independent cohorts. We performed clustering analysis to group CSF proteins into modules, each with similar sex and aging trajectories. Because aging plays a significant role in the pathophysiology of age-related diseases, we subsequently examined these modules for disease and gene ontology. Our findings show distinctive aging trajectories and sex differences in CSF proteome and their connection to common chronic diseases.

**Table 1.** The sex and aging distribution of the individuals included in this study.

| Healthy individuals | CSF |  |  | Plasma |
| --- | --- | --- | --- | --- |
|  | Knight-ADRC | ADNI | FACE | Knight-ADRC |
| **N** | 660 | 166 | 168 | 1,382 |
| **Males** | 280 (42%) | 89 (54%) | 83 (49%) | 565 (41%) |
| **Females** | 380 (58%) | 77 (46%) | 85 (51%) | 817 (59%) |
| **Age (mean, range)** | 70 (43, 91) | 75 (60, 90) | 66 (48, 86) | 73 (44, 91) |

## Results

### Sex and aging effects of CSF proteomics are distinctive, compared to plasma

To examine the impact of aging and sex differences on CSF proteome, we collected CSF samples from 660 cognitively healthy individuals in the Knight Alzheimer Disease Research Center (Knight ADRC) and subsequently quantified proteomic data through aptamer-based SOMAscan platform (SOMAscan7k)^21^. After a rigorous data processing and quality control, 7,006 aptamers targeting 6,162 human proteins remained and were utilized in the study. By performing regression analysis in this cohort, we identified 4,682 proteins significantly associated with age (including NEFL, GDF15, and MYL4; Fig.1a; Supplementary Table 1) and 1,587 proteins significantly associated with sex at FDR < 0.05 (including PUDP, MYL4, PZP, and APCS; Fig. 1b; Supplementary Table 2). There were 1,172 proteins affected by both age and sex (Fig. 1c). Among the age associated proteins, 3,037 proteins (64.9%) including NEFL, GDF15, and CFD increased with age, while 1,645 proteins (35.1%) including NEU1 and ID2 decreased with age (Fig. 1d). Among the sex associated proteins, 831 proteins (52.4%; including MYL4 and APCS) had higher abundance levels in males, while 756 proteins (47.6%; including PUDP and PZP) had higher abundance levels in females (Fig. 1d). The remaining 1,909 proteins such as BCHE and GSTT1 were not affected by either age or sex (Fig. 1d).

**Fig. 1:**
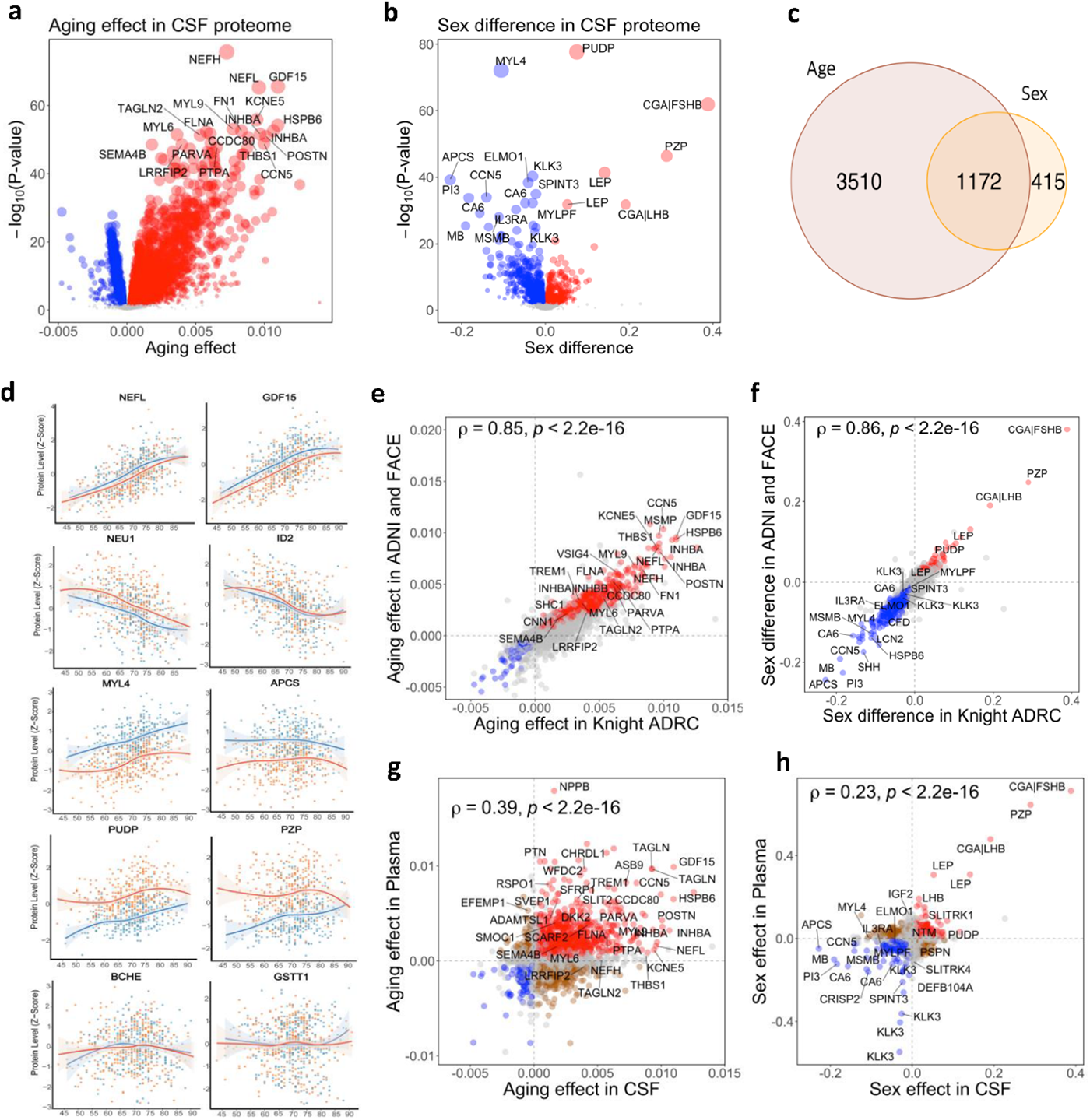
Age effects and sex differences in CSF proteomics. (a) Age effects on CSF proteins in Knight ADRC. (b) Sex differences in CSF proteins in Knight ADRC. (c) Significant proteins associated with age and/or sex at FDR < 0.05. (d) Protein abundance across ages for selected proteins in males (blue dots) and females (orange dots). The locally estimated scatterplot smoothing (LOESS) lines were separately in males (blue) and females (red). (e) Comparison of age effects on CSF proteins in independent ADNI and FACE (y-axis) versus those in Knight ADRC (x-axis). (f) Comparison of sex differences in CSF proteins in ADNI and FACE versus those in Knight ADRC. (g) Comparison between aging effects on plasma proteins versus those in CSF proteins. (h) Comparison between sex differences in plasma proteins versus those in CSF proteins

To validate these age and sex effects on CSF proteomics, we obtained the same SOMAscan7k proteomic data in CSF from 334 individuals in the Alzheimer’s Disease Neuroimaging Initiative (ADNI) and Fundació ACE Alzheimer Center Barcelona (FACE) cohorts, which were examined for aging effects and sex differences (Supplementary Tables 3-4). Out of the 4,682 age associated proteins in Knight ADRC, 1,942 proteins (41.5%) were nominally significant in this validation data at P < 0.05. Of the 1,587 sex associated proteins in Knight ADRC, 939 proteins (59.2%) were nominally significant in this data. When checked with all 7,006 analytes, correlation between the Knight ADRC and the validation data was very high (Pearson correlation ρ =0.85, *P* < 2.0×10^-16^, for aging effect, Fig 1c; ρ=0.86, *P* < 2.0×10^-16^, for sex difference; Fig. 1d), indicating the robustness of these identified aging effects and sex differences in CSF proteins.

To examine whether these findings would be specific to CSF or shared with plasma, we also obtained the SOMAscan7k proteomic data in plasma from 1,384 healthy individuals in Knight ADRC. In plasma data, we found 3,296 proteins associated with age (Supplementary Fig.1a; Supplementary Table 5) and 3,873 proteins associated with sex (Supplementary Fig.1b; Supplementary Table 6) at FDR < 0.05. There were 1,803 proteins associated with both age and sex (Supplementary Fig.1c). These age and sex effects on plasma proteome was also validated with the external UKB-PPP (ρ =0.44 for aging effect, Fig 1c; 2 for sex difference; p =0.5 Supplementary Fig. 1d-e). These moderate correction values were somewhat expected as the different proteomic platforms were used (SOMAscan in Knight ADRC vs. OLINK in UKB-PPP).

When these effects on plasma proteomics were compared to those on CSF proteomics, their correlations were much reduced (ρ=0.39 for aging effect, Fig 1c; ρ for sex difference; Fig. 1g-h). The aging effects of 1,824 (55.3%) proteins were not consistently identified in CSF and plasma, with 693 (21.0%) showing the opposite directions between CSF and plasma. While 1,472 (44.7%) proteins (including GDF15; β_A_=0.011 in CSF vs 0.0099 in plasma; Fig. 1g; Supplementary Fig. 1f) had the same directions in both CSF and plasma, several of these proteins had pronounced magnitude differences in their aging effect. For example, PTN increased with age 13 times faster in plasma than in CSF (β_A_ =0.0008 in CSF vs 0.0105 in plasma; Fig. 1g; Supplementary Fig. 1f), while MYL4 increased with age seven times faster in CSF than in plasma (β_A_ =0.0034 in CSF vs 0.0005 in plasma). With sex effects, only 502 (13.0%) proteins (including CGA|FSHB; βS =0.389 in CSF vs 0.713 in plasma; Supplementary Fig. 1f) had the same direction in CSF and plasma. For the remaining 3,371 (87.0%) proteins, sex effects were not consistent in CSF and plasma. For example, C3 in CSF had higher abundance in males, but C3 in plasma had higher abundance in females (βS =-0.125 in CSF vs 0.112 in plasma; Supplementary Fig. 1f). In contrasts, LANCL1 in CSF had higher abundance in females, but LANCL1 in plasma had higher abundance in males (βS =0.038 in CSF vs -0.054 in plasma; Fig. 1h; Supplementary Fig. 1f). These findings point that CSF proteome show distinctive age and sex differences compared to plasma proteome.

### CSF biological age is consistent to plasma biological age

To examine the biological age using proteomics in CSF and in plasma, we constructed a proteomic clock based on the 569 Knight ADRC samples that had proteomic data in both CSF and plasma. Since we found that proteins showed sex differences, we constructed sex-stratified biological clocks. We split the data into testing vs training (3/4 vs 1/4) and subsequently selected the model based on four-fold cross-validation. In CSF from males, the optimal proteomic aging model included 120 proteins (Fig. 2a; Supplementary Table 7), which provided the biological age highly consistent with the chronological age (correlation ρ=0.97 in training p =0.75 in testing data; Fig. 2b). Similarly, in CSF from females, the biological age predicted based on 134 proteins was highly consistent with the chronological age (ρ =0.97 in training data; ρ=0.87 in testing data; Fig. 2b). In the independent ADNI and FACE cohorts, these sex-specific biological clocks based on CSF proteomics provided comparable performance (in ADNI p=0.72 for males and ρ=0.61 for females; in FACE, ρ=0.75 for males and ρ=0.61 for females; Supplementary Fig. 2a). Compared to the CSF aging model, the proteomic aging model in plasma included fewer proteins (62 proteins in males and 89 proteins in females; Fig. 2a; Supplementary Table 7). The biological aging in plasma performed equally well (ρ=0.92 in training males; ρ=0.86 in testing males; ρ=0.94 in training females; ρ=0.83 in testing females; Fig. 2c). These plasma clocks were validated well in an additional set of 815 Knight-ADRC samples (ρ=0.83 for males and ρ=0.81 for females; Supplementary Fig. 2b).

**Fig. 2:**
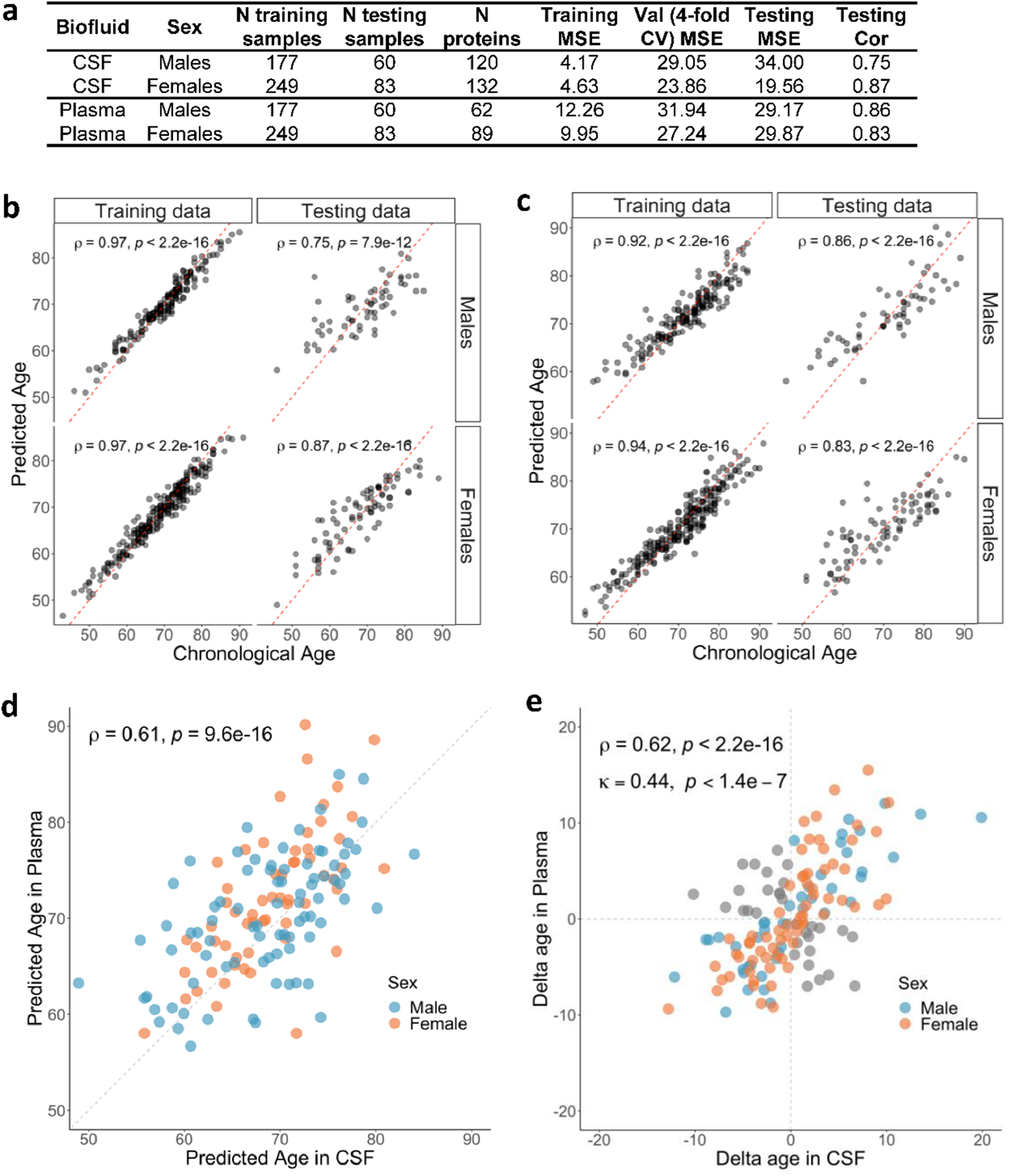
Sex-specific proteomic clocks in CSF and plasma. (a) Summary of creating sex-specific proteomic clocks. (b) Performance of CSF proteomic clock with chronological ages (x-axis) and predicated biological ages in CSF (y-axis). (c) Performance of plasma proteomic clock. (d) Comparison between CSF predicted age (x-axis) and plasma predicated age (y-axis) in males (blue) and females (red). (e) Comparison of delta ages in CSF versus in plasma. The delta age is a departure of the predicted age from the chronological age.

To compare the predicted biological age based on CSF or plasma clocks, we examined 143 Knight ADRC samples in which plasma and CSF were collected within 6 months. Across all samples, there was no statistically significant difference in the predicted biological ages based on CSF or plasma clocks (paired t-test P=0.915). The correlations between predicted biological age based on CSF or plasma clocks was moderate (ρ=0.61; Fig. 2d). When the correlation was examined separately for each sex, males showed somewhat higher correlation than females (ρ=0.67 in males; ρ=0.58 in females).

To examine whether predicted biological age based on CSF or plasma clocks had a similar relationship with chronological age (i.e., delayed, or accelerated aging), we computed the delta (the departure of each predicated biological age from the chronological age) in CSF and plasma. The overall correlation of the difference between predicted age based on CSF and plasma clocks and chronological age was ρ=0.620 with almost no difference in males and females (ρ=0.624 in males; ρ=0.632 in females; Fig. 2e). For 103 individuals (72%), the differences between age predicted by both CSF and plasma clocks and chronological age were in the same direction (kappa statistic=0.44). Fifty-three individuals (23 males; 30 females) showed delayed aging based on both the CSF and plasma clock, where predicted biological age was younger than the chronological age, while 50 samples (20 males; 30 females) showed accelerated aging with their biological age older than the chronological age. In the remaining 40 samples (28%; 17 males; 23 females), the differences between age predicted by both CSF and plasma clocks and chronological age were in different directions (e.g., the individual’s CSF clock provided accelerated aging, while his/her plasma clock provided delayed aging). Together, these results show that CSF proteomic data generally provide consistent information as the plasma proteomic data in terms of individual’s biological age.

### Network analyses of CSF proteomics identify modules with distinctive age and sex signatures

To reduce the complexity of the proteome, we performed weighted gene co-expression network analysis (WGCNA) with 7,006 aptamers in the Knight ADRC CSF samples. The network analysis provided 16 modules, which collectively included a total of 5,131 aptamers targeting 4,917 proteins (Fig 3a; Supplementary Table 8). The remaining 1,875 aptamers were not assigned into any of the 16 modules. When we visually examined each module with locally estimated scatterplot smoothing (LOESS) in males and females separately, each module included proteins with similar aging or sex signatures (Fig. 3b; Supplementary Fig. 3). Among them, several modules exhibited non-linear age trajectories.

**Fig. 3.**
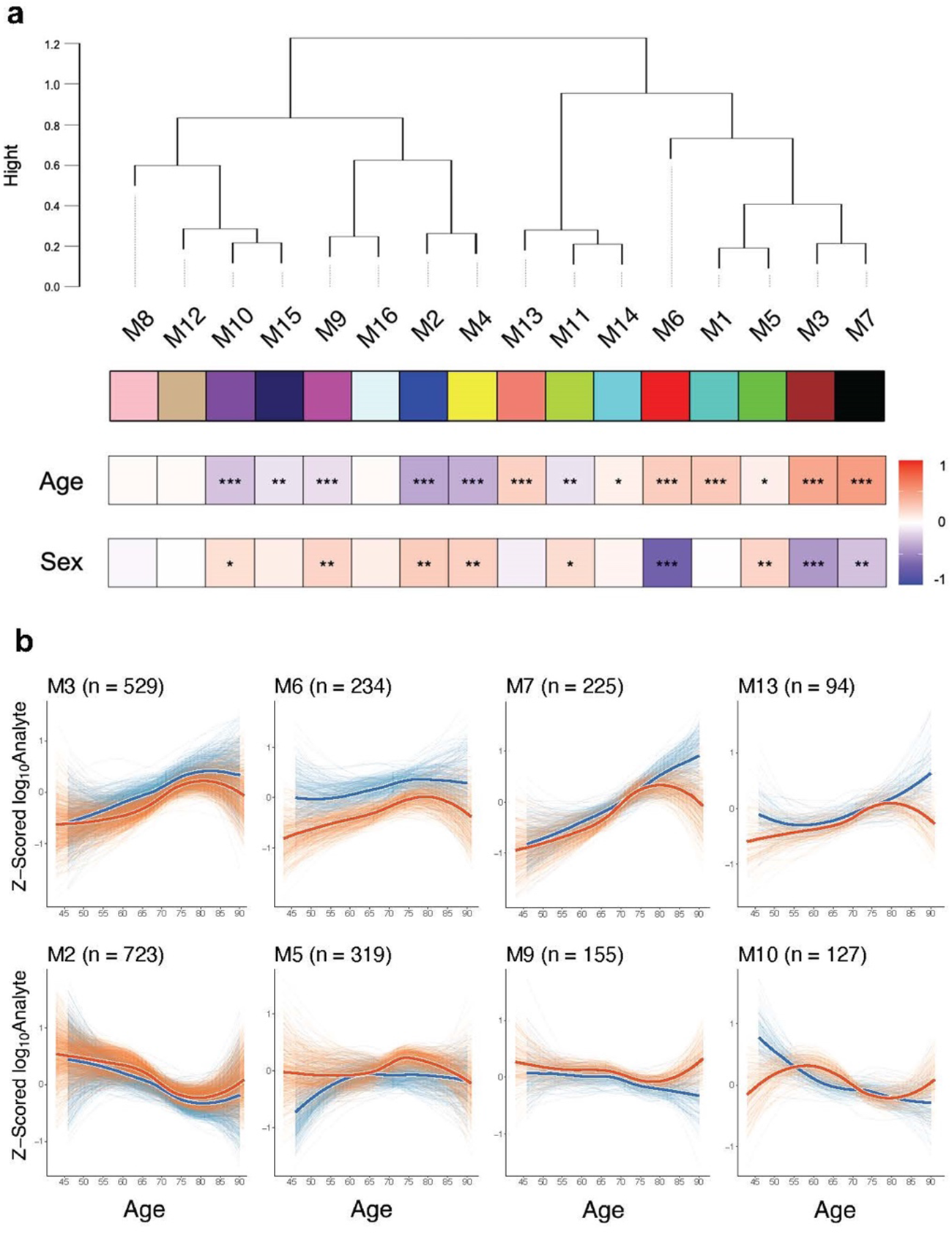
Clustering of CSF proteins into 16 modules using WGCNA. (a) WGCNA identified the 16 proteins modules. The age row shows the correlation coefficient between proteins and ages (red for increasing with age; blue for decreasing). The sex row shows Cohen’s distance between proteins in females vs those in males (red for higher in females; blue for higher in males). *** for *P* < 0.0001, ** for *P* < 0.001, and * for *P* < 0.05. (b) Aging trajectory for the select modules. LOESS lines for individual proteins in males (thin blue lines) and females (thin orange lines). The thick lines are the average across them separately in males (blue) and females (red). The number of included analytes (n) is shown. Aging trajectory for all modules is shown in Supplementary Fig. 3.

When each module was tested for aging effects, we found 13 modules significantly correlated with age (at *P*<0.05; Fig. 3a; Supplementary Table 9). Correlations within each sex (in males and females separately) were like those in the combined data (Supplementary Table 9). For some modules (M3, M4, and M6), aging trajectories in males and females were parallel with similar patterns (for example, showing protein levels increasing or decreasing around the same age). Several modules (including M10 and M15) had complex aging trajectories. For example, females increasing their protein abundances around the age when males decreasing their proteins. Seven modules (M1, M3, M5, M6, M7, M13, and M14) were positively correlated with age, showing increasing protein abundances as aging (Fig. 3a). Among them, M7 (including *KCNE5, HSPB6,* and *MYL9*) had the highest positive correlation (ρ=0.50, *P*=3.3×10^-42^). in M7 showed a stronger linear aging trajectory in males, whereas proteins showed non-linear aging trajectory with increasing abundance until 75 years old and decreasing afterward. In M5, the male trajectory increased until 65 years old and remained constant afterward, whereas its female trajectory stayed constant until 65 years old and changed afterward. With strong nonlinear trajectory, the correlation with age for M5 (ρ=0.082; *P*=3.45×10^-02^) did not capture full aging effect. Five modules (M2, M4, M9, M10, and M15) were negatively correlated with age, showing decreasing protein abundances as aging. M2 (including *NEU1* and *ID2*) had the lowest correlation, showing most decreases (ρ=-0.37, *P*=7.2×10^-23^).

When each module was tested for sex differences, nine modules showed statistically significant sex differences at *P* < 0.05 (Fig. 3). In three modules (M3, M6, and M7), males had higher protein levels. Among them, M6 (including *APCS, ELMO1, PI3, PLG*) showed the most noticeable sex difference with the largest Cohen’s distance (-0.70, *P*=1.8×10^-17^), whose aging trajectories in males and females were parallel. In six modules (M2, M4, M5, M10 and M11), females had higher protein levels. Among them, M2 (including *ENPP2, FIBP,* and *TNNI1*) showed the largest sex differences (Cohen’s d=0.25, P=1.51×10^-03^), with non-linear aging trajectories parallel in males and females.

### Enrichment analysis highlights modules linked to multiple aging diseases

To identify the protein modules relevant to common aging diseases, disease ontology enrichment was performed by examining enrichment for the diseases classified in the international classification of diseases’ 10^th^ revision (ICD-10). For the 1,145 diseases that were examined with each of 16 modules, 106 diseases were significantly enriched with eight modules (M2, M3, M5, M6, M7, M9, M13, and M15) at FDR < 0.05 (Fig. 4a; Supplementary Table 10). To uncover mechanistic insights into their relevance with diseases, we subsequently performed gene ontology (GO) enrichment for these eight modules. Six modules (M2, M3, M5, M6, M7, and M13) showed enrichments for GO biological processes (BP) at FDR < 0.05 (Supplementary Table 11). To examine cellular context of these modules and individual proteins, we also performed enrichment analysis using cell type expression data from human brain cells^22^ (Supplementary Table 12).

**Fig. 4:**
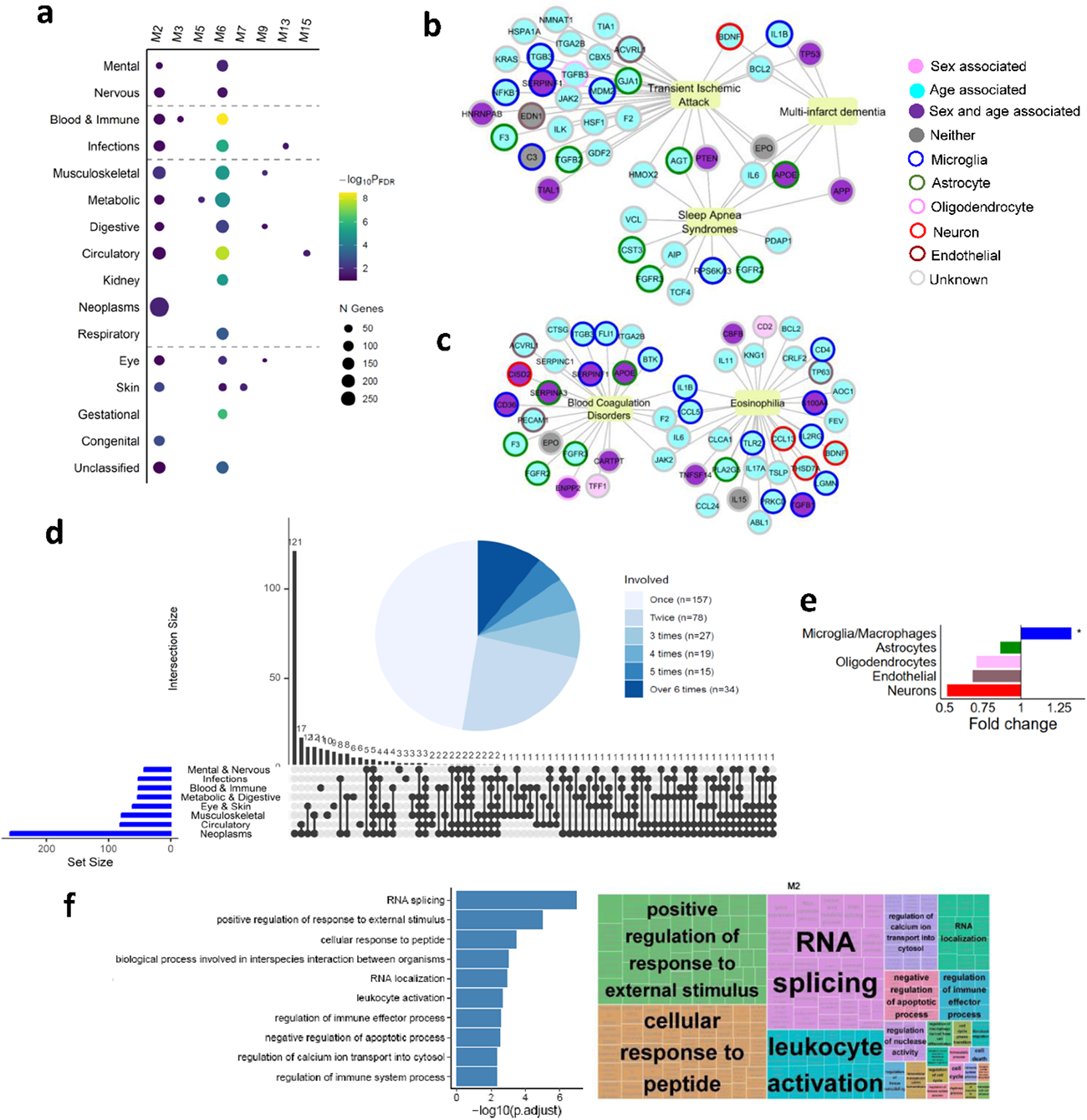
Enrichment of module M2. (a) The enrichment of 8 modules for the 16 ICD-10 disease chapters. (b) M2 proteins involved with mental and neuro-diseases. (c) M2 proteins involved with blood and immune mechanisms. (d) Upset plot for M2 proteins enriched with the 11 chapters. Congenital and unclassified are excluded. The pie chart shows the number of M2 proteins overlapping across the ICD-10 chapters (e) Cell-type specificity of M2 (f) Gene ontology enrichment of M2.

Six modules (M3, M5, M7, M9, M13 and M15) showed enrichment for three or fewer IDC-10 chapters. M3 showed enrichment for the complement deficiency disease in the blood and immune disease chapter (FDR = 2.42×10^-02^), involving 11 proteins including *C7, C4A, C1S,* and *C3* (Supplementary Fig. 4a), mainly expressed in the microglia and astrocytes. M3 showed enrichment for 8 GO pathways including extracellular matrix organization (FDR = 2.55×10^-02^) and complement activation (FDR = 2.55×10^-02^), explaining its link to complement deficiency disease (Supplementary Fig. 4b). M5 was enriched for lysosomal storage diseases, mucopolysaccharidoses (FDR=6.89×10^-03^), involving seven proteins including IDUA, NT5E, GNS and EGFR (Supplementary Fig. 4c). They were enriched for 25 GO pathways including synapse assembly (FDR = 7.09×10^-03^) and the lysosomal protein catabolic process (FDR = 2.67×10^-02^; Supplementary Fig. 4d). M7 was enriched for the keloid skin disease (FDR = 8.79×10^-03^) involving 15 proteins including TAGLN, SERPINE1, and FLNA (Supplementary Fig. 4e). They showed enrichment for GO pathways in actin cytoskeleton organization (FDR = 6.88×10^-06^), which regulates several essential wound-healing components^23^, thereby explaining its link to skin diseases (Supplementary Fig. 4f). M9 was enriched for the musculoskeletal system (spondylitis, FDR = 4.45×10^-03^), the digestive system (recurrent aphthous ulcer, FDR = 0.02), and eye diseases (keratoconjunctivitis, FDR = 0.026), involving multiple proteins including IFNG, PLG and IL23R (Supplementary Fig. 4g). M13 was enriched for enterovirus infections (FDR = 1.55×10^-02^) involving eight proteins (VIM, STAT1, and G6PD; Supplementary Fig. 4h). They were enriched for GO pathways involved in translation (FDR = 2.84×10^-02^) and stress granule assembly (FDR = 4.99×10^-02^; Supplementary Fig. 4i). Stress granules formed from mRNAs modulates the stress response and viral infection^24^, explaining its link to infections and parasitic diseases. M15 was enriched for dilated cardiomyopathy (FDR=1.05×10^-02^), with ten proteins including CASP8, GCH1, TNNT2, and ADCYAP1, mainly expressed in the microglia and neuron (Supplementary Fig. 4j).

### Two modules are linked to neurodegeneration and multiple aging diseases

We found that modules M2 and M6 were enriched for multiple ICD-10 chapters, suggesting their relevance to comorbidity conditions across multiple aging diseases (Supplementary Table 10).

M2 showed significant enrichment for 32 diseases spanning 13 ICD-10 chapters at FDR < 0.05 (Fig. 4a). 43 proteins including *APOE*, *APP,* and *IL6* were involved with mental disorders (multi-infarct dementia, FDR = 0.029) and diseases in the nervous system (transient ischemic attack, FDR=2.96×10^-02^; sleep apnea syndrome, FDR = 3.44×10^-02^; Fig 4b). Enrichment for blood and immune mechanism was found with 52 proteins including JAK2, CCL5, and IL1B; eosinophilia, FDR = 0.029; blood coagulation disorders, FDR = 4.20×10^-02^; Fig: 5c). Enrichment for circulatory system was notable (81 proteins including APOE, AGT, SERPINA3; subarachnoid hemorrhage, FDR = 2.96×10^-02^; cerebral amyloid angiopathy, FDR = 2.96×10^-02^; Supplementary Fig. 5a). M2 was the only module that was enriched for neoplasms (acute megakaryocytic leukemias, FDR = 8.74×10^-03^; prostate cancer, FDR =1.52×10^-02^), involving 259 proteins including CD36 and ANGPTL4 (Supplementary Fig. 5b). Proteins involved with the additional six ICD-10 chapters were presented in Supplementary Fig. 6. Proteins in M2 were enriched for microglia and macrophage (57 proteins including NFKB, C3, and IL1B, enrichment P = 1.34×10^-02^, Fig. 4e).

For 384 GO pathways, belonging to the 25 higher-level representative terms, M2 showed enrichment (Fig. 4f). Pathways related to neurodegeneration and immune mechanisms include cellular response to peptide (response to stress, FDR = 2.63×10^-04^; neuron apoptotic process, FDR = 2.19×10^-02^) and negative regulation of apoptotic process (FDR = 3.16×10^-03^). Several pathways related to neoplasm were found, including apoptosis, leukocyte activation (FDR=2.54×10^-03^), cell cycles and cell death (necrosis), RNA splicing (FDR = 1.08×10^-07^), and RNA localization (FDR=1.20×10^-02^). Cancer cells exploit RNA splicing to promote tumor growth^25–28^. When tumor cells avoid apoptosis, regulated cell death is activated during tumor cell proliferation in defense tumor progression and migration^29,30^. Adhesion cascade leads to localization of inflammatory cells to tumor sites including the recruitment of effector leukocytes^31^. In addition, pathways involved leukocyte activation explain this module’s relevance to peripheral T-cell lymphoma and other types of leukemia.

Among all 16 modules, M6 showed enrichment for the largest number of diseases (70 belonging to 14 ICD-10 chapters) with the highest statistical significance (Fig. 4a; Fig. 5a). In M6, 61 proteins including LRRK2, SERPINA3, and C3 were involved with mental disorders (mild cognitive disorder, FDR = 8.37×10^-03^; schizophrenia, FDR = 2.51×10^-02^) and nervous system (multiple sclerosis, FDR = 1.81×10^-02^; late onset AD, FDR = 5.02×10^-02^; Fig. 5b). The most significant enrichment was found for blood and immune mechanism (complement deficiency disease, FDR = 2.14×10^-09^; blood coagulation disorders, FDR = 7.96×10^-08^), involving 68 proteins including PLG, SELE, SERPINA3 (Fig. 5c). In addition, the circulatory system (cardiovascular diseases, FDR = 1.16×10^-08^; atherosclerosis, FDR = 4.81×10^-07^; Supplementary Fig. 7a), infections (septicemia, FDR = 1.65×10^-06^; Supplementary Fig. 7b), metabolic diseases (diabetes mellitus, FDR = 1.04×10^-05^; Supplementary Fig. 7c) were strongly enriched for this module. Proteins involved with the additional five ICD-10 chapters were presented in Supplementary Fig. 8.

**Fig. 5:**
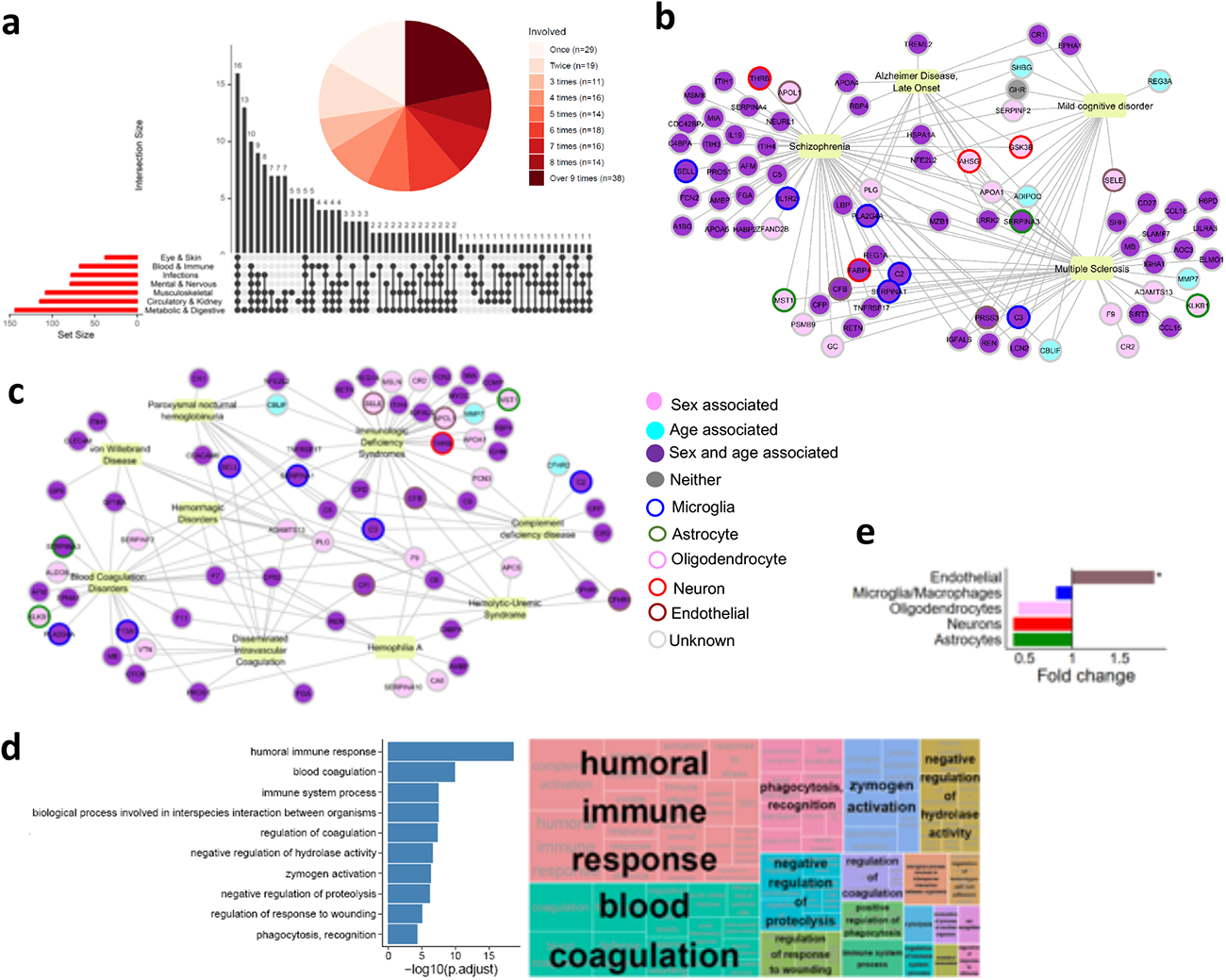
Enrichment of module M6. (a) Upset plot for M6 proteins enriched with the 11 chapters. Respiratory, congenital and unclassified were excluded. The pie chart shows the number of M6 proteins overlapping across the ICD-10 chapters. (b) M2 proteins involved with mental and neuro-diseases. (c) M2 proteins involved with blood and immune mechanisms. (d) Gene ontology enrichment of M6 (e) Cell-type specificity of M6.

Among all 16 modules, M6 also had the highest statistical significance for GO pathways, showing enrichment for 126 GO biological processes belonging to the 18 higher-level representative terms at FDR < 0.05 (Fig. 5d). The strongest enrichment was involved with humoral immune response (FDR=2.14×10^-19^; complement activation, FDR = 2.14×10^-19^). In particular, complement activation plays a defensive role in our body through proteolytic cascades and opsonization^32^, explaining a direct link to complement deficiency disease. The second strongest enrichment involved blood coagulation (FDR=5.64×10^-10^; coagulation, FDR = 1.189×10^-10^; defense response, FDR = 2.47×10^-08^), which explains the link to blood coagulation disorders of the blood and immune system. Other pathways included phagocytosis recognition (FDR = 6.13×10^-05^), zymogen activation (FDR = 5.73×10^-07^), and regulation of coagulation (FDR = 5.89×10^-08^). Proteins in M6 were enriched for endothelial cells (15 proteins including APOL1 and SELE; enrichment P = 1.41×10^-02^, Fig. 5e).

## Discussion

In this study, we systematically investigated the aging effects and sex differences on CSF proteomics in a large scale. CSF protein levels can be influenced only by factors or diseases that influence the brain and the central nervous system, showing more direct relevance to brain aging and diseases. By comprehensively examining 7,006 aptamers targeting 6,139 proteins in CSF obtained from 660 healthy individuals aged from 43 to 91 years old, we subsequently identified significant sex and aging effects on 5,097 aptamers in CSF. They were confirmed by two additional independent cohorts. We found that many of the sex and aging effects on CSF proteins had different magnitude or even opposite direction, when compared to their corresponding effects on plasma proteins, indicating distinctive CSF-specific signatures. Clustering of these CSF proteins revealed not only the modules with clear aging trajectories but also the modules with sex differences in their protein abundance. We subsequently identified the relevance for complex diseases for the eight modules, out of which two modules (M2 and M6) showed striking connection to neuropsychiatric diseases and many aging diseases.

In this study, we identified significant age-associated increase in GDF15, NEFH, NEFL, and IL6. The growth differentiation factor 15 (GDF15) is one of the transforming growth factor-beta cytokine superfamily essential in regulating cellular response to stress signals. Elevated GDF15 levels are reported be associated with pathological conditions involving inflammation, myocardial ischemia, and cancer^33,34^. Increase of GDF15 with aging has been reported in several studies^16^, including their link to mitochondrial function decline with aging^35^. Neurofilament heavy polypeptide (NEFH) is one of type IV intermediate cytoplasmic filament proteins in neurons^36^, which provide structural support for the axon and regulate axon diameter, thus maintaining the transmission of electrical impulses along axons^37^. Neurofilament light polypeptide (NEFL) is a neuronal cytoplasmic protein from the member of the intermediate filament protein family, highly expressed in myelinated axons. It increases in CSF and blood along with axonal damage in neurological disorders including inflammatory, neurodegenerative, traumatic and cerebrovascular diseases^38–40^. Age-associated increase of NEFL has been previously reported^41–43^. Interleukin 6 (IL6), a multifunctional cytokine, is a major contributor to acute phase inflammatory response. While it is normally present in low levels, IL6 is increased in aging and with chronic age-related diseases including neurodegeneration, diabetes, rheumatoid arthritis, cancer, atherosclerosis, and infections^44–46^. In addition, this study confirmed several of 82 age-associated proteins (including PTN, MB, and CST3) identified by Baird et al^47^. Johnson et al in his systemic review compiled 32 age-associated proteins that were consistently reported across multiple studies^16^. Out of these 32 proteins, our study confirmed the significant aging effects of 23 proteins including GDF15, ANXA1, C3, C4A, and EGFR^16,17,48–52^ in both CSF and plasma.

This study found the strong sex difference in APOE, SERPINA3, C3, PLG, SERFINF2, SELE, APCS, which are known to be critical for multiple diseases including neurodegeneration, cardiovascular disease (CVD), kidney failures, and diabetes. Apolipoprotein E (APOE), a carrier protein for cholesterol and lipids, is produced in abundance in the brain and serves as the principal lipid transport vehicle in CSF^53^. It is critically involved in the pathogenesis of AD and CVD, through induction at high concentration in peripheral nerve injury and repair by redistribution of lipids to regenerating axons and to Schwann cells during remyelination. The APOE gene alleles modulate human aging^53–56^. SERPINA3, a part of protease inhibitors, is critically involved in the anti-inflammatory response and antiviral responses^57^, and identified as a specific biomarker of delirium and AD^58,59^. Complement component 3 (C3) is the central point of the three-cascade activation pathway of the complement system. The complement plays a role in inflammatory processes, metabolism, apoptosis, mitochondrial function, Wnt signaling pathways, and plays a significant role in aging-related diseases, including AD, macular degeneration, and osteoarthritis^60,61^. Plasminogen (PLG) is the zymogen of plasmin, a broad specificity serine protease whose activity contributes to pathological conditions including cardiovascular diseases, infections, metabolic disorders, asthma, kidney disorders, and musculoskeletal diseases^62–68^. SERPINF2 is a serine protease inhibitor of plasmin, modulates insulin sensitivity, and is associated with CVD, blood coagulation disorders, rheumatoid arthritis, asthma, and diabetes^69–71^. Soluble endothelial leukocyte adhesion molecule-1 (SELE), a part of cell adhesion molecules expressed only on endothelial cells by cytokines, is a pro-inflammatory protein important in cell-cell/cell-matrix interactions during immune responses and inflammatory process^72^. Elevated levels are associated with CVDs, diabetes, cerebrovascular diseases and rheumatoid arthritis^73–76^. Serum amyloid P component (APCS) is a soluble pattern-recognition protein, binding to damaged membranes, nuclear autoantigens, and determinants on microbial pathogens and recruiting other elements of host defense^77^. APCS is involved in the regulation of matrix formation, regulation of complement activation and an acute phase response protein in response to infection and inflammatory cytokines during the innate immune response^78^

Multiple studies including Lehallier et al. clustered plasma proteins into modules and found non-linear changes throughout the lifespan, which may be important to fully appreciate the undulating changes^17^. In this study, we examined the clustering of the CSF proteins into modules and identified not only the modules with often non-linear aging trajectories but also the modules with clear sex differences. Module 6 showed the strongest sex differences and the most extensive enrichment for multiple aging diseases including neurodegeneration. Several studies demonstrated the immune system’s role in contribution to diverse pathological conditions, which aligns with our findings. Immune system functions have been associated with neurodegeneration including AD. Gate et al. revealed the adaptive immune response in AD^79^ and demonstrated the T-cell involvement in Lewy body dementia^80^. In addition, we found strong connection of two module (M2 and M6) to complement deficiency disease. Complement, a group of over 30 proteins involved in the complement pathway or cascade, is critical in the inflammation response of the innate immune system against micro-organisms and infections^81–86^. The complement system orchestrates opsonization, facilitate cytotoxic destruction, formulate membrane attack complexes (MAC), and the liberation of peptides that promote inflammatory responses^86^. Complement deficiencies arise when any of these proteins are missing from the cascade or fail to function properly, which results in increase prevalence in infections by pyogenic organisms via phagocytosis^81,86^. Both modules showed enrichment of the related biological processes involved in translation, phagocytosis, stress granule assembly, and cell-cell adhesion^87–96^.

Several clocks based on transcriptomics, methylations, or proteomics in plasma provided insights for biological aging and their relevance to the diseases. These studies demonstrated that biological aging is profoundly involved in the pathogenesis of many diseases and age-related health conditions. In this study, we constructed sex-specific proteomic clock in both CSF and plasma. While an individual protein may change differently in CSF and plasma as aging, the performance of the CSF or plasma-based clocks was similar in terms of the correlation with the chronological age. Furthermore, they also provided similar inference on whether an individual has delayed or accelerated aging. These results indicate that brain aging may be like the overall biological aging based on circulating plasma proteomics.

Our study has some limitations. First, while this study has the largest sample size to examine aging and sex differences in CSF proteomics from healthy individuals as of today, it is based on a cross-sectional study. Therefore, a longitudinal follow-up study may be needed to validate the findings from this study. Second, while the aging and sex effects of CSF proteins were often different from those of plasma proteins, the sex specific proteomic clock in these biofluid provided similar performance. This may be because this study examined only healthy individuals, who often exhibit a less pronounced degree of accelerated aging, when compared to individuals with diseases. An additional follow-up will be needed to distinguish the organ-specific biological aging. Third, to examine the relevance of CSF proteins to biological processes and diseases, we employed statistical and computational approaches. Additional follow-up in the select proteins will be needed for their clinical utility.

In summary, we comprehensively examined age effects and sex differences in over 6,000 CSF proteins, identifying 4,682 age-associated and 1,587 sex-different proteins. These age effects and sex differences were distinctive of CSF, when compared to plasma. Many of these proteins were involved in multiple diseases including neurodegeneration and cardiovascular diseases. Our findings based on CSF proteomics fill a gap of current aging research and help understanding the role of age and sex in CSF protein regulation during a healthy lifespan. They may help developing personalized treatment for aging interventions and clinically translatable interventions for brain aging and diseases.

## Methods

### Study populations

This study examined 1,807 cognitively normal individuals from the Knight Alzheimer Disease Research Center (Knight-ADRC)^97–99^, Alzheimer’s Disease Neuroimaging Initiative (ADNI)^100^, and Fundació ACE (FACE) ^101^ studies (Table 1). The age of these individuals spanned from 43 to 91 years old, representing middle and elderly adults. The Knight-ADRC at Washington University in St. Louis aims to advance AD research with the goal of treatment or prevention of AD. The ADNI is a longitudinal multicenter study designed to develop clinical, imaging, genetic, and biochemical biomarkers for the early detection and tracking of AD. Headquartered in Barcelona, FACE has collected and analyzed almost 18,000 genetic samples, diagnosed over 8,000 patients. This study was approved by the Institutional Review Boards of the Washington University School of Medicine in St. Louis, and the research was performed in accordance with the approved protocols.

### Proteomics data

Cerebrospinal fluid (CSF) samples from the Knight-ADRC, ADNI, and FACE were collected using lumbar puncture (LP) in the morning after an overnight fast. All samples underwent the identical protocols for preparation and processing, and stored at -80 °C. All samples were randomized across plates to avoid batch effects and were sent together to SomaLogic. Protein levels were quantified using the SOMAscan platform based on a multiplexed, single-stranded DNA aptamer assay. The quantitative levels of 7,584 aptamers were reported as relative fluorescence unit (RFU). Initial data normalization was performed by SomaLogic using hybridization controls for intra-plate and median signal to account for inter-plate variances^102^. Normalization against an external reference to control for biological variances was also performed by SomaLogic^103^. In-house quality control (QC) was also performed to exclude outliers. More details of the QC process were published elsewhere^103,104^. After processing, CSF proteomic data consisted of 7,006 aptamers targeting 6,139 proteins in CSF from 994 individuals. Plasma proteomic data consisted of 6,905 aptamers targeting 6,106 proteins in plasma from 1,372 samples.

### Age effects and sex differences in CSF and plasma proteomes

To examine the effect of age and sex on CSF protein abundance, we performed regression analysis using the following model:

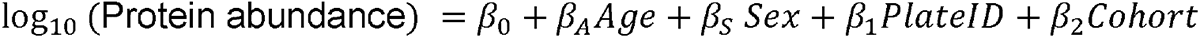

Our primary interest is the age effect β_A_ and the sex effect β_S_ on log_10_-transformed protein abundance. Age is an individual’s age at LP; sex is coded as 1 for a female and 0 for a male; PlateID is a plate identifier for proteomic sample; and Cohort is a cohort information (Knight-ADRC, ADNI, FACE). Analysis was performed first using the Knight-ADRC data, where a cohort was excluded in the model. Additional analysis was performed using the combined set of ADNI and FACE data. To control for multiple testing, the false discovery rate (FDR) < 0.05 based on the FDR-BH method was used.^105^ In addition, to compare the effect of sex and age on CSF proteomics with those of plasma proteome, the same analysis was performed using the Knight-ADRC plasma proteomic data.

### Proteomics based biological clock

We constructed sex-stratified predictive models for biological age using proteomic measurements with the least absolute shrinkage and selection operator (LASSO) regression. Four models were constructed: aging clocks for males and females, each separately with CSF proteomics and plasma proteomics. To compare the predictive models between CSF and plasma proteomic data, we considered 569 individuals that had both CSF and plasma samples. We used log transformed RFU values of 6,738 analytes in both CSF and plasma proteomic data. We split the data into training data (3/4 with 177 males and 249 females) and testing data (the remaining 1/4 with 60 males and 83 females). The R package glmnet^106^ was used for LASSO regression analysis. The shrinkage parameter that controls the model complexity was determined using 4-fold cross-validation on the training data. To examine model performance, we examined Pearson correlation coefficient and root mean squared error (RMSE) between the predicted ages and the actual chronological age.

### Network analyses of CSF proteomics

To cluster CSF proteins into modules, we performed the weighted gene co-expression network analysis (WGCNA)^107^. Specifically, we utilized the *blockwiseModules function* with the following parameters: power=7, corType=” bicor”, networkType=” signed”, deepSplit=4, reassignThreshold=0.05, and mergeCutHeight=0.15. After adjusting for covariates, the soft threshold power was determined as 7 by considering both the scale-free topology model fit (R^2^=0.9595) and mean connectivity. We used bicor, a median-based measure of similarity that is more robust to outliers than the Pearson or Spearman correlation coefficient^108^. A signed network was considered to account for both positive and negative correlations. The topological overlap matrix (TOM) was first calculated to determine the interconnection within the network structure. Hierarchical clustering analysis was conducted based on the calculated 1-TOM values to classify the proteins into distinct modules. The final modules were defined by grouping and rearranging similar groups using the dynamic tree cut method^109^. For each module, we calculated the first eigenvector, known as the eigen protein. For the aging effect in each module, we considered the Pearson correlation coefficient between the eigen protein and age. For the sex differences in each module, we considered Cohen’s distance^110^ between male eigen proteins and female eigen proteins.

### Sex-specific aging trajectory

To visualize aging trajectories of each protein, we used the locally estimated scatterplot smoothing (LOESS)^111^ method in R. To examine sex differences, we drew LOESS lines for males and females, separately. In addition, to examine the sex-stratified aging trajectories in each module, we obtained the average line of LOESS lines in males and females, separately, of all the proteins belonging to that module. In addition, to determine whether these sex-stratified aging trajectories in the Knight-ADRC data were reproducible, we examined proteomic data from two independent data (ADNI and FACE) in the same manner. Specifically, we created a grid point for age and calculated the Pearson correlation coefficients of the LOESS values at these points between the two datasets. We performed a one-sided correlation test at the FDR < 0.05 using the FDR-BH method^105^. Only those validated proteins were included in the subsequent enrichment analysis.

### Enrichment analysis

For disease ontology, we performed enrichment analysis using the enrichDGN function in the R package DOSE^112^. The Entrez gene names for the validated proteins in each module were considered. To convert the CUI^113^ codes used by DisGeNET into the international classification of disease, 10th revision (ICD-10) codes developed by World Health Organization, we used the Python package Owlready2 (ref^114^). ICD-10 codes^115^ are available at https://icd.who.int/browse10/2019/en. Using the first three digits of ICD-10 codes, 1,145 diseases were grouped into 22 chapters. Cases without a matching ICD code were excluded from the analysis.

For Gene Ontology (GO) biological processes, we performed enrichment analysis using enrichGO function in the R package clusterProfiler^116^. Significant GO terms were visualized into the hierarchically related GO pathways based on their similarity to distill them into common higher-level representative terms using REVIGO^117^. GO IDs were restricted to Homo sapiens species and visualization was created with treemaps.

To obtain cellular context of these modules and individual proteins, we also performed enrichment analysis using cell type expression data from human brain cells^22^ (Supplementary Table 12). Based on the cell type data from gene expression data from human astrocytes, neurons, oligodendrocytes, microglia/macrophages, and endothelial cells, we obtained the degree of specificity to relevant cell types for the proteins. For 5,750 proteins, these cell-type expression data were available. Fisher’s exact test was used to examine cell-type enrichment of each module against the cell-types for 5,750 proteins.

## Data Availability

All data generated in this study are included in this published article.

## Declarations

### Ethics approval and consent to participate

Ethics approval for every individual cohort was obtained from the respective Institutional Review Boards and research was carried out in accordance with the approved protocols (WUSTL IRB approval 201109148). Written informed consent was obtained from participants or their family members and all participating institutions approved the study design.

### Consent for publication

All authors have approved the contents of this manuscript and provided consent for publication.

### Availability of data and materials

All data generated in this study are included in this published article.

### Funding and Acknowledgements

We thank all the participants and their families, as well as the involved cohorts, institutions, and their staff.

This work was supported by grants from the National Institutes of Health, RF1 AG074007 (YJS), R01 AG044546 (CC), P01 AG003991 (CC), RF1 AG053303 (CC), RF1 AG058501 (CC), U01 AG058922 (CC), P30 AG066444 (JCM), P01 AG003991 (JCM), P01 AG026276 (JCM), and the Chan Zuckerberg Initiative (CZI), the Michael J. Fox Foundation (CC), and the Alzheimer’s Association Zenith Fellows Award (ZEN-22-848604, awarded to CC).

<u>ADNI acknowledgement:</u> Data collection and sharing for this project was funded by the Alzheimer’s Disease Neuroimaging Initiative (ADNI) (National Institutes of Health Grant U01 AG024904) and DOD ADNI (Department of Defense award number W81XWH-12-2-0012). ADNI is funded by the National Institute on Aging, the National Institute of Biomedical Imaging and Bioengineering, and through generous contributions from the following: AbbVie, Alzheimer’s Association; Alzheimer’s Drug Discovery Foundation; Araclon Biotech; BioClinica, Inc.; Biogen; Bristol-Myers Squibb Company; CereSpir, Inc.; Cogstate; Eisai Inc.; Elan Pharmaceuticals, Inc.; Eli Lilly and Company; EuroImmun; F. Hoffmann-La Roche Ltd and its affiliated company Genentech, Inc.; Fujirebio; GE Healthcare; IXICO Ltd.; Janssen Alzheimer Immunotherapy Research & Development, LLC.; Johnson & Johnson Pharmaceutical Research & Development LLC.; Lumosity; Lundbeck; Merck & Co., Inc.; Meso Scale Diagnostics, LLC.; NeuroRx Research; Neurotrack Technologies; Novartis Pharmaceuticals Corporation; Pfizer Inc.; Piramal Imaging; Servier; Takeda Pharmaceutical Company; and Transition Therapeutics. The Canadian Institutes of Health Research is providing funds to support ADNI clinical sites in Canada. Private sector contributions are facilitated by the Foundation for the National Institutes of Health (www.fnih.org). The grantee organization is the Northern California Institute for Research and Education, and the study is coordinated by the Alzheimer’s Therapeutic Research Institute at the University of Southern California. ADNI data are disseminated by the Laboratory for Neuro Imaging at the University of Southern California.

### Author’s contributions

DS, CML and CA performed analysis, interpreted the results, and drafted the manuscript. GH contributed to the manuscript preparation. JT performed proteomics data processing and quality control. PK acquired Knight ADRC samples and data. MB, AO, MVF and AG acquired phenotypes and CSF samples in FACE. JCM and SES obtained funding, recruited Knight ADRC cohort, and curated phenotype data. TSP obtained funding, supervised the work, and interpreted the results. YJS and CC obtained funding to generate proteomic data, designed the study, supervised the work, interpreted the results, and drafted the manuscript. All authors read and approved the manuscript.

